# Spatio-temporal distributions of COVID-19 vaccine doses uptake in the Netherlands: A Bayesian ecological modelling analysis

**DOI:** 10.1101/2023.03.09.23287033

**Authors:** Haoyi Wang, Tugce Varol, Thomas Gültzow, Hanne M. L. Zimmermann, Robert A.C. Ruiter, Kai J. Jonas

## Abstract

**Background:** In the transitioning era towards the COVID-19 endemic, there is still a sizable population that has never been vaccinated against COVID-19 in the Netherlands. To identify regions and populations that have a lower chance of vaccination uptake, this study provides a spatio-temporal estimation of the relative chance of COVID-19 vaccination uptake for the first, second, and the booster doses in the Netherlands on both municipality level and the public health services (regional) level.

**Methods:** Data on COVID-19 vaccination uptake were retrieved from the publicly available national COVID-19 surveillance dataset. We used a Bayesian spatio-temporal modelling technique with the integrated nested Laplace approximation to account for the spatial structure and the space-time interaction. Additionally, we used an ecological regression modelling technique which takes into account areal level socio-demographic characteristics to adjust for their potential impact on the chance of the regional vaccination uptake.

**Results:** Our findings revealed a heterogenous spatio-temporal distribution of the relative chance of COVID-19 vaccination uptake with highly overlapping trends of all three vaccination doses. Internal heterogeneity of COVID-19 vaccination uptake within one public health services region on the municipality level was also identified. The Dutch main urban area and the most religiously conservative regions were identified to have a lower-than-average chance of COVID-19 vaccination uptake compared to the rest of the country. Ecological regression modelling analysis revealed that regions with a higher proportion of non-Western immigrants had a lower chance of COVID-19 vaccination uptake for all vaccination scenarios.

**Conclusion:** The obtained estimates should inform national and local COVID-19 vaccination policies and service strategies in the Netherlands for the ongoing COVID-19 campaign on the second booster. Namely, more regional efforts and services may be needed to close ‘vaccination gaps’ and optimise COVID-19 health-related outcomes, especially with regard to regions with a relatively higher proportion of marginalised populations.

## Introduction

The coronavirus disease 2019 (COVID-19) caused by Severe Acute Respiratory Syndrome Coronavirus 2 (SARS-CoV-2) was declared a pandemic by the World Health Organization (WHO) in March 2020 [1] and is now transitioning towards an endemic status in many countries [2, 3], including the Netherlands. One of the reasons for the ‘way out of the pandemic’ was due to the fast-developed COVID-19 vaccines [2, 4].

To facilitate COVID-19 vaccination uptake in the Netherlands, great efforts had been made by the Dutch National Institute for Public Health and the Environment (RIVM), through promoting public campaigns [5, 6] and conducting behavioural studies to inform public health communication and policies [7]. Unfortunately, a sizeable part (16.5%) of the Dutch adult population still have not get vaccinated against COVID-19 according to the latest available data as of September 4, 2022 [5, 8]. Additionally, vaccine hesitancy was reported to be present in the Netherlands [9-11], which the World Health Organisation (WHO) listed as one of the ten global threats to health [12].

Several determinants have been reported to be associated with individuals’ vaccine hesitancy from previous studies, e.g., safety concerns and side effects of vaccines, a distrust of COVID-19 vaccines and governments, having a low socio-demographic position, e.g., due to belonging to a ethnic minority groups, holding a low level of education, or being financially disadvantaged [13-15]. Most of these insights focus on individual risk profiles and individuals’ beliefs underlying their decision-making for vaccination uptake. Yet, to further close ‘vaccination gaps’ to prevent serious COVID-19-related health outcomes, evidence from an ecological perspective is warranted, too, such as identification of the geographical and temporal clusters of regions on a small area level and populations with a lower chance of vaccination uptake. With this information COVID-19 vaccine-related services and interventions can be better tailored and targeted to populations with a lower chance of vaccination uptake with higher needs [4].

To investigate COVID-19 vaccination uptake on a small area level and to provide robust estimations Bayesian spatio-temporal analysis can be used. Bayesian spatio-temporal analysis is a well-established method for small-area-estimations [16-21]. Briefly, Bayesian spatio-temporal analysis can account for several sources of error or bias including spatial autocorrelation between neighbouring regions and proximity and time-dependent autocorrelation between consecutive periods in sparsely populated areas, compared to the observed frequentist prevalence calculation [20-22]. Despite Bayesian spatial and spatio-temporal analysis having been applied to monitor the distribution of COVID-19 infections in some countries, such as the United States [22], and the distribution of COVID-19 vaccination uptake, such as in Belgium [23], it has not been applied in the Netherlands for COVID-19 infections nor for vaccination uptake monitoring. Statistical approaches that have been used in the Netherlands stays at the frequentist calculation [8].

Therefore, in this study, we aimed to apply Bayesian spatio-temporal analysis to identify clusters of lower COVID-19 vaccination uptake, among its population on both the municipality and the public health services (in Dutch: Gemeentelijke of Gemeenschappelijke Gezondheidsdienst [GGD], which is a smaller regional health-specific administrative level) levels in the Netherlands. As a secondary objective, we investigated whether indentifying clusters of lower COVID-19 vaccination uptake on a finer-defined geographical unit would reveal a more precise or different spatio-temporal pattern of COVID-19 vaccination uptake. Also, given the established evidence on how socio-demographic factors can influence individuals’ vaccination COVID-19 decisions [13-15], we aimed to explore how these socio-demographic factors may impact the overall COVID-19 vaccination uptake on an ecological level.

## Material and methods

### Study population

#### Study area

In the Netherlands, the planning, monitoring, and evaluation of public health measures, including COVID-19 vaccination, are mostly carried out by the GGDs. The Netherlands has 25 GGD regions in total. Within the GGD regions, the smallest administrative units are on the municipality level, which entails 345 municipalities in total. Estimates on both GGD and municipality levels provide valuable information for Dutch policymakers [20].

#### Data sources

We retrieved surveillance data on COVID-19 vaccination uptake, by vaccination scenarios, from RIVM with the openly accessible COVID-19 vaccination uptake data [5]. For the beginning phase of the vaccination promotion, due to the requirement of privacy protection by RIVM, data on the vaccination prevalence in regions with less than 5% coverage were masked. We, assumed a 0% coverage for these regions.

For the socio-demographic spatial proxies, we retrieved freely accessible data per municipality/GGD from Statistics Netherlands (CBS), which provides reliable statistical information across the Netherlands to produce insight into social issues [24], and linked them with the COVID-19 vaccination uptake surveillance data on the areal level. The following datasets were used in this study: a) “Bevolking 15 tot 75 jaar; opleidingsniveau, wijken en buurten” for data on proportion of low education, 2019 [25]; b) “Bevolking; migratieachtergrond, generatie, leeftijd, regio, 1 januari, 2021” for data on proportion of non-western migration background population [26]; and c) “Kerncijfers wijken en buurten 2021” for data on the proportion of financially extremely disadvantaged population [27].

#### Study population and vaccination scenarios

We included data of all populations who were 18 years and older in this study. In the Netherlands, five different vaccines have been used: Moderna (Spikevax), BioNTech/Pfizer (Comirnaty), AstraZeneca (Vaxzevria), Janssen, and Novavax [5]. Given that the majority of the administrated COVID-19 vaccines were the 2-doses based mRNA COVID-19 vaccines, such as Pfizer/BioNTech and Moderna [28], we conducted analyses on three different COVID-19 vaccination scenarios (hereinafter vaccination scenarios), namely 1) covered primary partly (only one dose of the selected vaccine has been administered), 2) covered primary completed (two doses of the selected vaccine have been administered), and 3) covered first booster (covered primary completed and one booster dose), following the definitions and terminologies from RIVM [5]. Only a minority of the Dutch population has been vaccinated against COVID-19 using the 1-dose-based viral vector COVID-19 vaccine, Janssen [28]. For this population, following RIVM’s recommendation, we considered administrating one dose as primarily completed [29].

#### Study periods

The first COVID-19 vaccines were administered on January 6, 2021, in the first-dose vaccination promotion in the Netherlands [5]. After the first-dose vaccination promotion, the second-dose promotion and the booster-dose proportion were started in February 2021 and November 2021, respectively. To achieve maximum vaccination uptake [6], in the Dutch vaccination program, populations with clinical health vulnerabilities were vaccinated first, followed by stratification based on age groups from the elderly to the young. Given the different vaccination-promoting periods, to avoid misleading spatio-temporal interaction estimation, we only retrieved the periods when the COVID-19 vaccination was available to our total selected study population. As a result, we only included data on COVID-19 vaccination covered primary partly from February 2021 to August 2022 (19 months in total); on COVID-19 vaccination covered primary completed from March 2021 to August 2022 (18 months in total), and on COVID-19 vaccination covered first booster from November 2021 to August 2022 (10 months in total).

### Bayesian spatio-temporal analysis

To estimate the relative chance of COVID-19 vaccination uptake on the small area levels (municipality and GGD levels), we used the Integrated Nested Laplace Approximation (INLA), which is a computationally less intensive but efficient and equivalent alternative to Markov chain Monte Carlo for the Bayesian computation [16, 17]. We appointed a Penalized Complexity (PC) prior for the precision of the exchangeable random effects [30], by employing the re-parameterised Besag-York-Mollie (BYM2) model [30]. This model specifies the spatially structured residual using an intrinsic conditional autoregressive distribution [30, 31], and uncertainties due to instability of estimates in sparsely populated areas [16, 18, 19].

In this study, we first applied this method to describe the spatio-temporal relative chance of COVID-19 vaccination uptake by the vaccination scenarios and two geographical levels with a space-time interaction to explore the spatio-tempotal trends. To further understand the relative chance of COVID-19 vaccination doses uptake in each selected period, we also investigated the spatial relative chance of COVID-19 vaccination uptake by the vaccination scenarios and two geographical levels over the selected periods without the space-time interaction (results in Supplementary materials S7 to S12).

### Spatio-temporal ecological modelling

In this study, we also explored whether COVID-19 vaccination uptake in the Netherlands was influenced by the selected regional socio-demographic characteristics as the spatial proxies on an ecological level. We, therefore, applied a spatio-temporal ecological regression modelling technique [16] which takes these spatial proxies into account to pick up additional associations and noises [21]. For the proportion of non-Western immigrants and the proportion of financially extremely disadvantaged individuals. data were retrieved from 2021, assuming these two spatial proxies were stable for the selected periods in this study. For the variable of the proportion of financially extremely disadvantaged individuals, we used data on people who were taking Bijstandsuitkering, a subsidy that is only available for those who are financially extremely disadvantaged. For data on the proportion of individuals with a low level of education, we extrapolated data from 2019, given that the most recent data was only available from 2019 from CBS. We assumed that the proportion of individuals with a low level of education from 2019 is comparable to 2021-2022 and stable over time.

We first conducted univariable models which only included one of the selected spatial proxies and the space-time interaction. The models’ goodness of fit was assessed using the deviance information criterion (DIC). We then conducted multivariable models with all determinants that were shown to significant in the univariable models based on Bayesian credible intervals (CrIs) and DIC, to evaluate the explained variance of COVID-19 vaccination uptake. CrIs can be regarded as a Bayesian analogue to confidence intervals to present the 95% probability of the posterior means [32]. All results on the spatio-temporal distribution of COVID-19 vaccination uptake presented in this study were based on the spatio-temporal final regression models.

### Model assumptions and parameter appointments for the spatio-temporal analysis of the relative chance of COVID-19 vaccination uptake in the Netherlands

In this study, for the counted outcome of vaccination uptake, we assumed that the observed vaccinated cases follow a Poisson distribution with mean *E*_*i*_ *θ* _*i*_, following the suggestions from Diggle (1983) on spatial analysis for disease mapping [33], where *E*_*i*_ is the expected number and *θ* _*i*_ is the relative chance in each region *i*. We thus can define the posterior relative chance on the logarithmic scale:

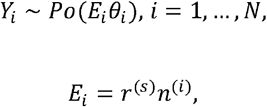

where *r*^(*s*)^ is the prevalence of COVID-19 vaccination uptake in the Netherlands, and *n*^(*i*)^ is the size of the population of region *i*. Additionally, we added a temporal dimension into the model with a spatio-temporal interaction, so the model was formed as:

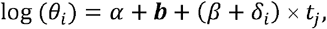

where (*β* + *δ*_*i*_) × *t*_*j*_ *stands* for the space-time interaction term:

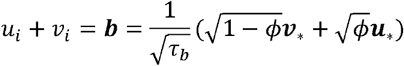

Where *α* represents the overall chance of the vaccination uptake in the Netherlands, *ui* is a random effect on area *i* which is used to model spatial dependence between the relative chance, and *vi* represents other unstructured noise which follows a distribution of 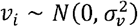. *τ*_*b*_ > 0 is a marginal precision parameter contribution from spatial term ***u***_*_ and random effect ***v***_*_, and the fraction of this variance explained by the from spatial term ***u***_*_ and random effect ***v***_*_ are the mixing parameter 0 ≤ *ϕ* ≤ 1.

In spatio-temporal ecological analysis, we first applied the univariable models which only include one of the selected determinants:

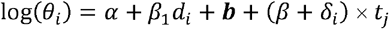

where *β*_1_ is the coefficient for the one of the determinants *d*_*i*_ selected in this study. In the final model, we included all the significant determinants indicated by the univariate models:

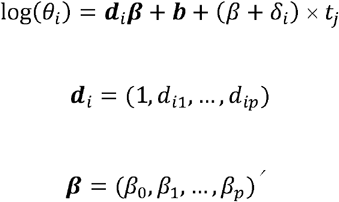

To define the PC prior, we used the probability statement 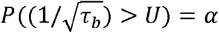. Based on the rule of thumb by Simpson et al. [30], we set *U* = 0.5/0.31 and *a* = 0.01. We then defined the prior for the mixing parameter *ϕ* as *P* (*ϕ* < 0.5) = 2/3, which assumed that the unstructured random effect accounts for more of the variability than the spatially structured effects [18].

All analyses were conducted in R (version 4.2.1). We used SpatialEpi R package (version 1.2.7) to estimate the expected COVID-19 vaccination uptake number per region [34]. For all Bayesian modelling analyses with INLA, we used the R-INLA package (version 21.05.02) [16].

### Ethical statement

The study only used publicly accessible secondary data from the national public surveillance system. Ethical approval and informed consent were therefore not required, given secondary data were depersonalised. Data was presented in an aggregated manner excluding the possibility of identifying specific individuals.

## Results

### Spatio-temporal distribution of COVID-19 vaccination uptake in the Netherlands

#### COVID-19 vaccination covered primary partly uptake, February 2021 to August 2022

Figure 2 presents the spatio-temporal distribution of the relative chance of COVID-19 vaccination uptake covered primary partly on the a) municipality level and b) GGD level, compared to the average chance of vaccination uptake on the national level, from February 2021 to August 2022.

**Figure 1.**
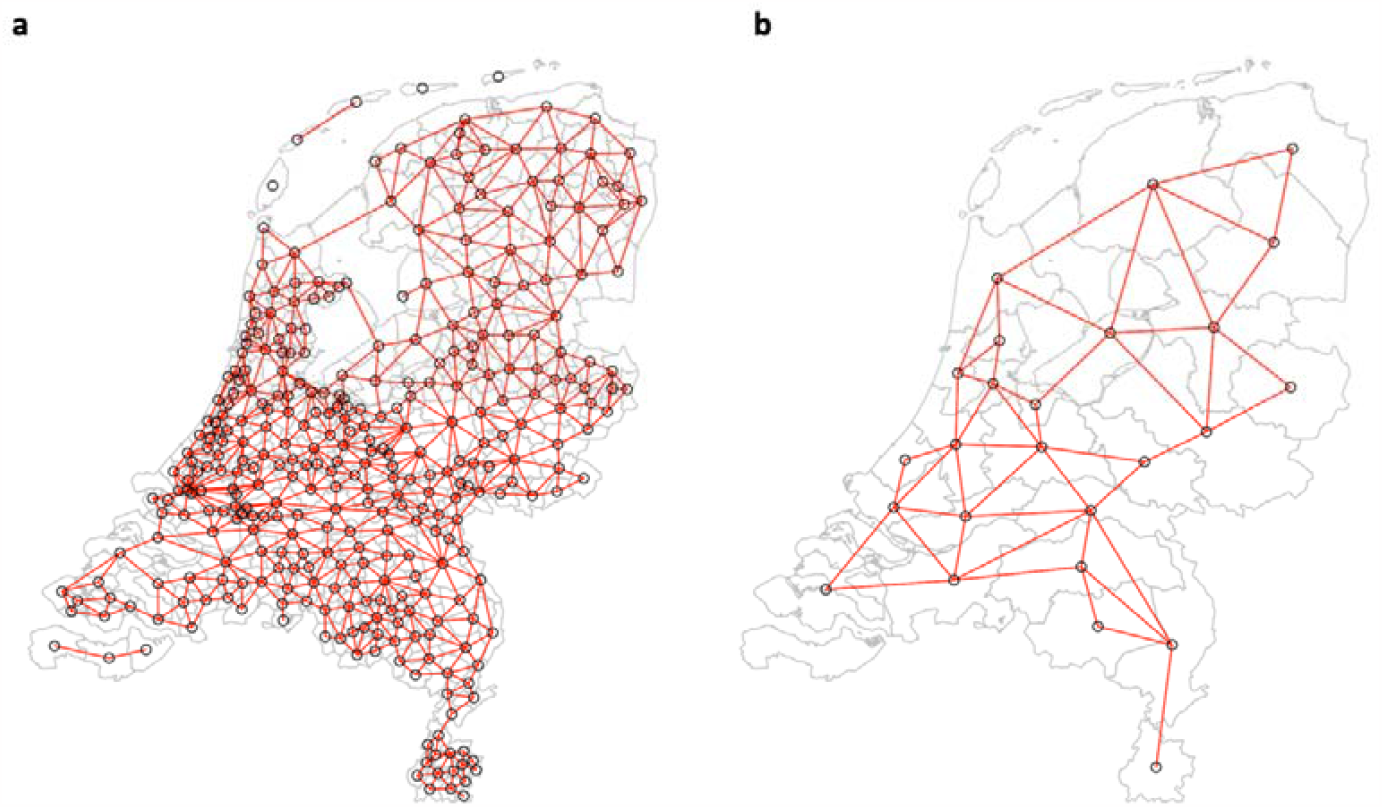
The Netherlands spatial connectivity between a) municipality level, and b) public health services (GGD) level. Notes: for names and more details on the municipalities in the Netherlands please see: https://www.government.nl/topics/municipalities; and Public Health Services regions in the Netherlands, please see: https://www.ggd.nl/.

**Figure 2.**
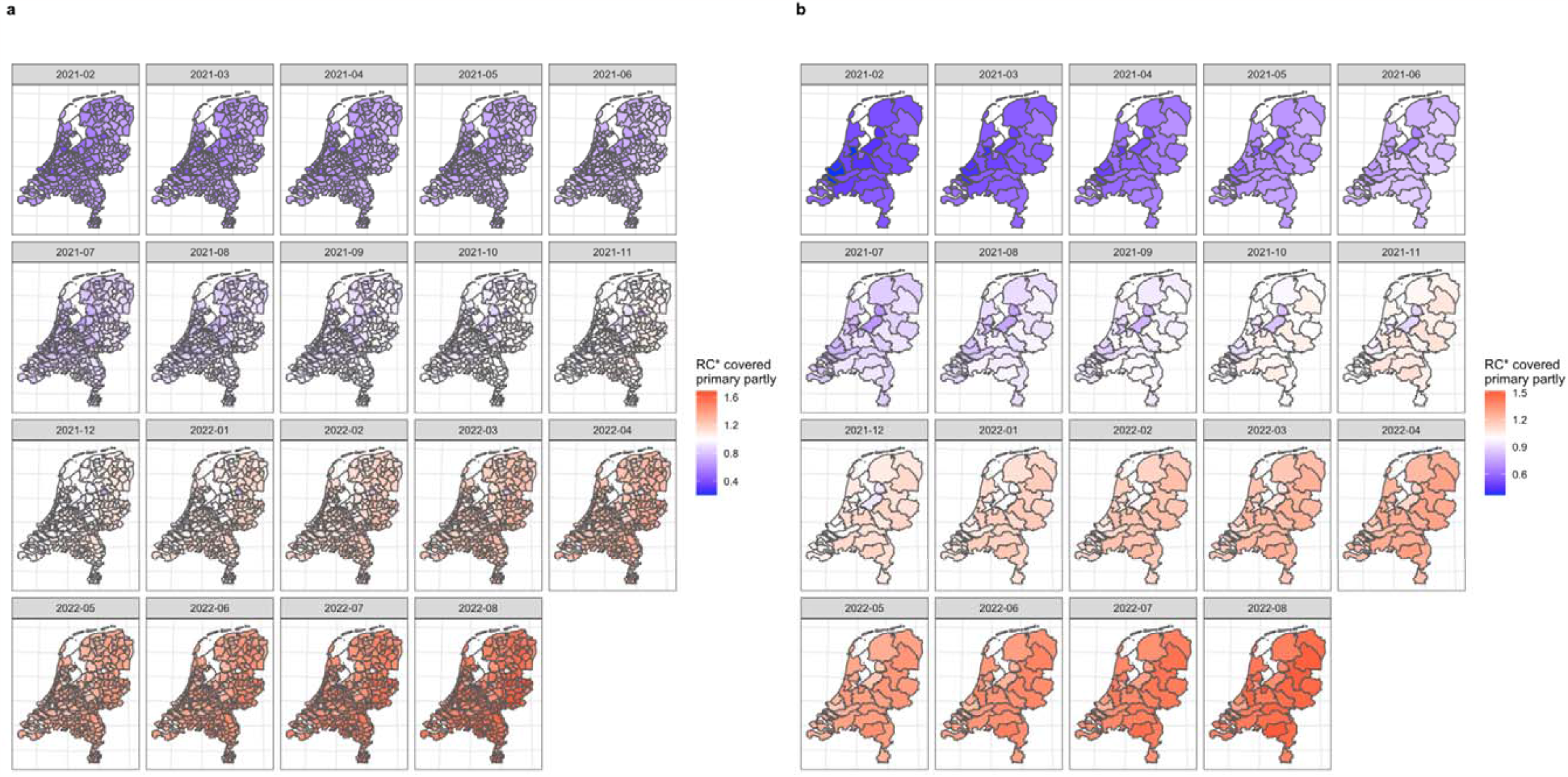
Choropleth map of the relative chance of COVID-19 vaccination uptake covered primary partly (only one dose administrated) on a) municipality level and b) Public health services (GGD) level by Bayesian spatio-temporal ecological modelling (final model), February 2021 to August 2022. Note: RC= relative chance, * indicates results estimated by Bayesian spatio-temporal ecological final model. For a better visibility, larger figures can be found in Supplementary materials S1 to S2. RC higher than 1 indicates a higher-than-average average risk in the Netherlands) chance of COVID-19 vaccination in that region (red); RC lower than 1 indicates a lower-than-average chance of COVID-19 vaccination in that region (blue).

Of the included periods, we identified a similar spatio-temporal trend of the COVID-19 vaccination uptake covered primary partly on both municipality and GGD levels. In general, of each selected temporal period, the highest relative chance of the COVID-19 vaccination uptake covered primary partly was found in the East of the Netherlands, while the lowest relative chances were found in the West, especially among municipalities close to Amsterdam and Rotterdam.

We also identified internal heterogeneity of the relative chances among municipalities within the GGD level over time. Taking the GGD Amsterdam region as an example, the overall relative chance of the COVID-19 vaccination uptake covered primary partly in February 2021 was estimated to be 0.368 (95% CrI 0.367;0.269). Within the GGD Amsterdam region, the relative chance among municipalities ranged from 0.374 (0.373;0.375) in Amsterdam to 0.614 (0.607;0.621) in Ouder-Amstel; and the overall relative chance in August 2022 was estimated to be 1.316 (1.314;1.318). The relative chance among municipalities ranged from 1.317 (1.314;1.319) in Amsterdam to 1.560 (1.551;1.569) in Aalsmeer.

#### COVID-19 vaccination covered primary completed, March 2021 to August 2022

Figure 3 presents the spatio-temporal distribution of the relative chance of the COVID-19 vaccination uptake covered primary completed on both a) municipality level and b) GGD levels, compared to the average chance of vaccination uptake on the national level from March 2021 to August 2022.

**Figure 3.**
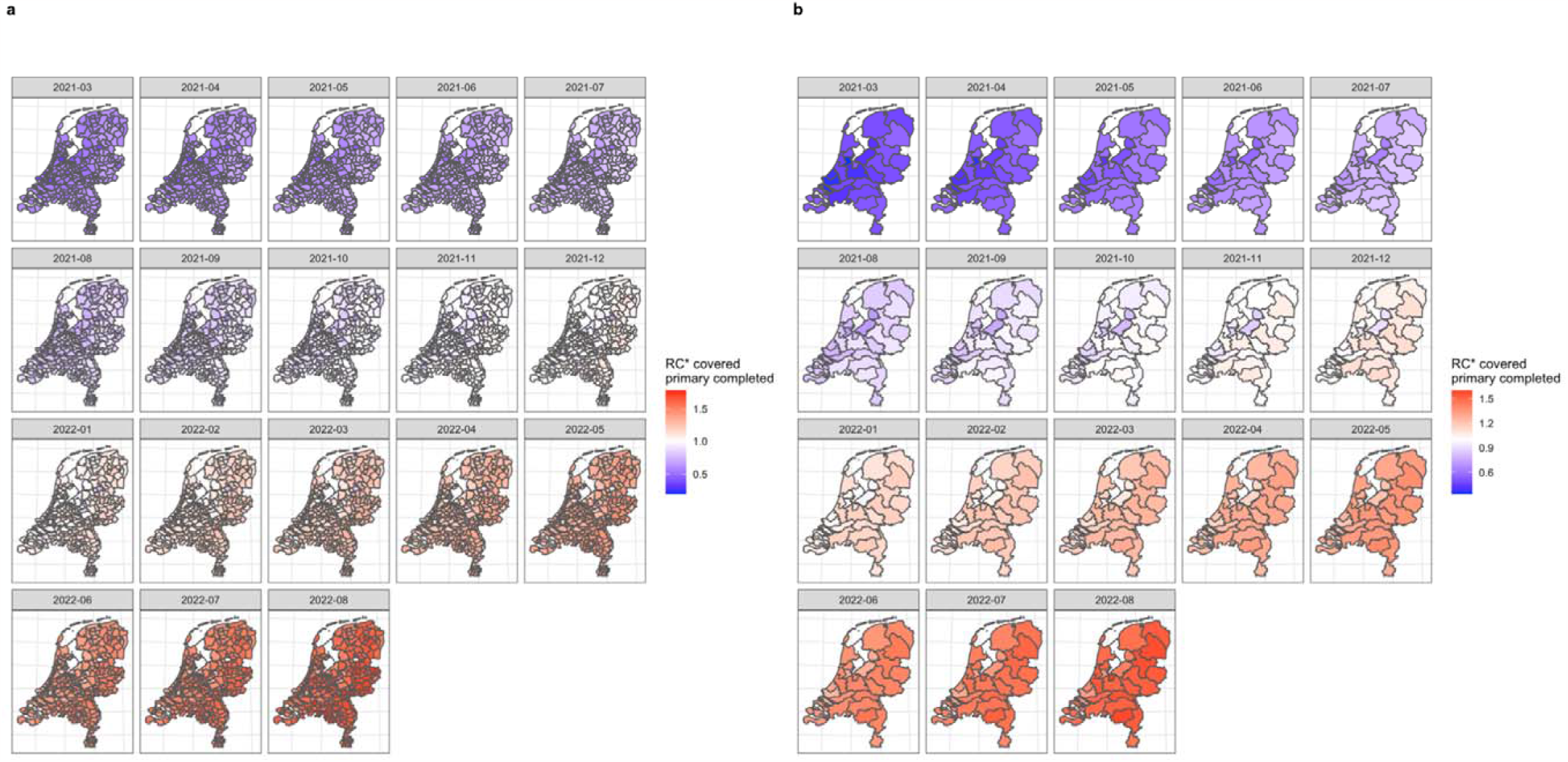
Choropleth map of the relative chance of COVID-19 vaccination uptake covered primary completed (two doses administrated) on a) municipality level and b) Public health services (GGD) level by Bayesian spatio-temporal ecological modelling (final model), March 2021 to August 2022. Note: RC= relative chance, * indicates results estimated by the Bayesian spatio-temporal ecological final model. For better visibility, larger figures can be found in Supplementary materials S3 to S4. RC higher than 1 indicates a higher-than-average (average risk in the Netherlands) chance of COVID-19 vaccination in that region (red); RC lower than 1 indicates a lower-than-average chance of COVID-19 vaccination in that region (blue).

We identified similar spatio-temporal trends of the relative chance on both geographical levels of the COVID-19 vaccination uptake covered primary completed as the spatio-temporal chances of the COVID-19 vaccination uptake covered primary completed with the spatial internal heterogeneity within the GGD regions. What is noteworthy to mention is, in the median period of November 2021, lower relative chances of the COVID-19 vaccination uptake covered primary completed were identified in regions from the Southwest to Northeast (in Dutch: Bible belt, this region is considered the most religiously conservative region in the Netherlands.)

#### COVID-19 vaccination covered first booster, November 2021 to August 2022

Figure 4 presented the spatio-temporal distribution of the relative chance of the COVID-19 vaccination uptake covered first booster on both a) municipality level and b) GGD levels, compared to the average chance of vaccination uptake on the national level and from November 2021 to August 2022.

**Figure 4.**
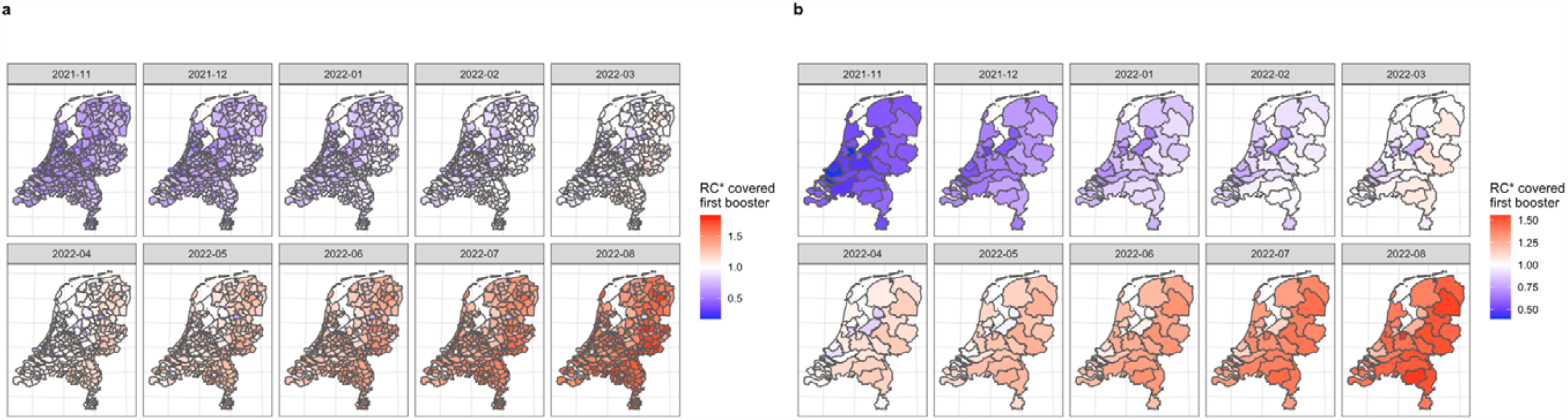
Choropleth map of the relative chance of COVID-19 vaccination uptake covered first booster (one booster dose administrated) on a) municipality level and b) Public health services (GGD) level by Bayesian spatio-temporal ecological modelling (final model), November 2021 to August 2022. Note: RC= relative chance, * indicates results estimated by the Bayesian spatio-temporal ecological final model. For better visibility, larger figures can be found in Supplementary materials S5 to S6. RC higher than 1 indicates a higher-than-average (average risk in the Netherlands) chance of COVID-19 vaccination in that region (red); RC lower than 1 indicates a lower-than-average chance of COVID-19 vaccination in that region (blue).

Again, similar spatio-temporal trends with internal spatial heterogeneity within the GGD regions were identified compared to the COVID-19 vaccination uptake covered primary partly and completed. Regions located around Amsterdam and Rotterdam, and those located in the Bible Belt, were again showing a lower relative chance of the COVID-19 vaccination uptake covered first booster compared to other regions in the Netherlands over the selected periods.

In addition, in this vaccination scenario, we found that, on GGD regional level, the GGD-Zuid-Limburg region, for example, showed a lower relative chance of the COVID-19 vaccination uptake covered first booster over the selected periods as well. Taking the median selected period of March 2021 as an example, the overall relative chance of the COVID-19 vaccination uptake covered first booster was estimated to be 0.953 (0.951;0.955), which can be considered as a significantly lower relative chance compared to the average national chance over the selected periods. While zooming in on the municipality level, the relative chances ranged from 0.726 (0.718;0.734) in Vaals to 1.148 (1.141;1.154) in Eijsden-Margraten which presented a significantly higher chance compared to the average chance of vaccination uptake on the national level.

For detailed relative chance per region and per selected period of each vaccination scenario on both geographical levels, see Supplementary Excel File 1. For the spatial distribution of the relative chance per region and per selected period of each vaccination scenario, see Supplementary materials S7 to S12.

### Bayesian ecological modelling on spatial socio-demographic proxies on COVID-19 vaccination uptake by COVID-19 vaccination scenarios

#### Univariable models

All detailed regional spatial socio-demographic proxies’ characteristics can be found in Supplementary Excel File 2. Univariably, for the three COVID-19 vaccination uptake scenarios, a higher proportion of non-Western immigrants and a higher proportion of financially extremely disadvantaged individuals were associated with a lower chance of COVID-19 vaccination uptake on both geographical levels significantly. Taking the COVID-19 vaccination uptake covered first booster on the municipality level as an example, for each one per cent increase of the proportion of non-Western immigrants in one municipality, the relative chance of the booster COVID-19 vaccination uptake in that municipality decreased by 0.9% (= 1-exp(−0.985*0.01)) over the selected periods. The proportion of people with a low level of education was only significantly associated with the COVID-19 vaccination uptake covered first booster on the municipality level (coefficient=-0.396, 95%CrI −0.666;-0.126), indicating that for each one per cent increase of the proportion of people with a low level of education in one municipality, the relative chance of the booster COVID-19 vaccination in that municipality decreased by 0.4% (= 1-exp(−0.396*0.01)) over the selected periods. This effect disappeared on the GGD level (coefficient=-0.085, −1.444;1.615).

#### Multivariable models

After adjusting for all the significant spatial socio-demographic proxies identified in the univariable model, multivariably, only the proportion of non-Western immigrants was found to be significantly negatively associated with the relative chance of COVID-19 vaccination uptake of all three vaccination scenarios on both geographical levels. Taking the COVID-19 vaccination uptake covered first booster on the municipality level as an example, for each one per cent increase of the proportion of non-Western immigrants in one municipality, the relative chance of the booster COVID-19 vaccination in that municipality decreased by0.9% over the selected periods.

For all univariable and multivariable models, we identified significantly increasing temporal trends. Taking the COVID-19 vaccination uptake covered first booster on the municipality level as an example, the relative chance of the first booster COVID-19 vaccination uptake increased by 9.7% (=exp(0.093) on average over the selected periods. For all detailed results obtained from other uni-/multivariable models for all three vaccination scenarios on both geographical levels, see Table 1.

**Table 1.**
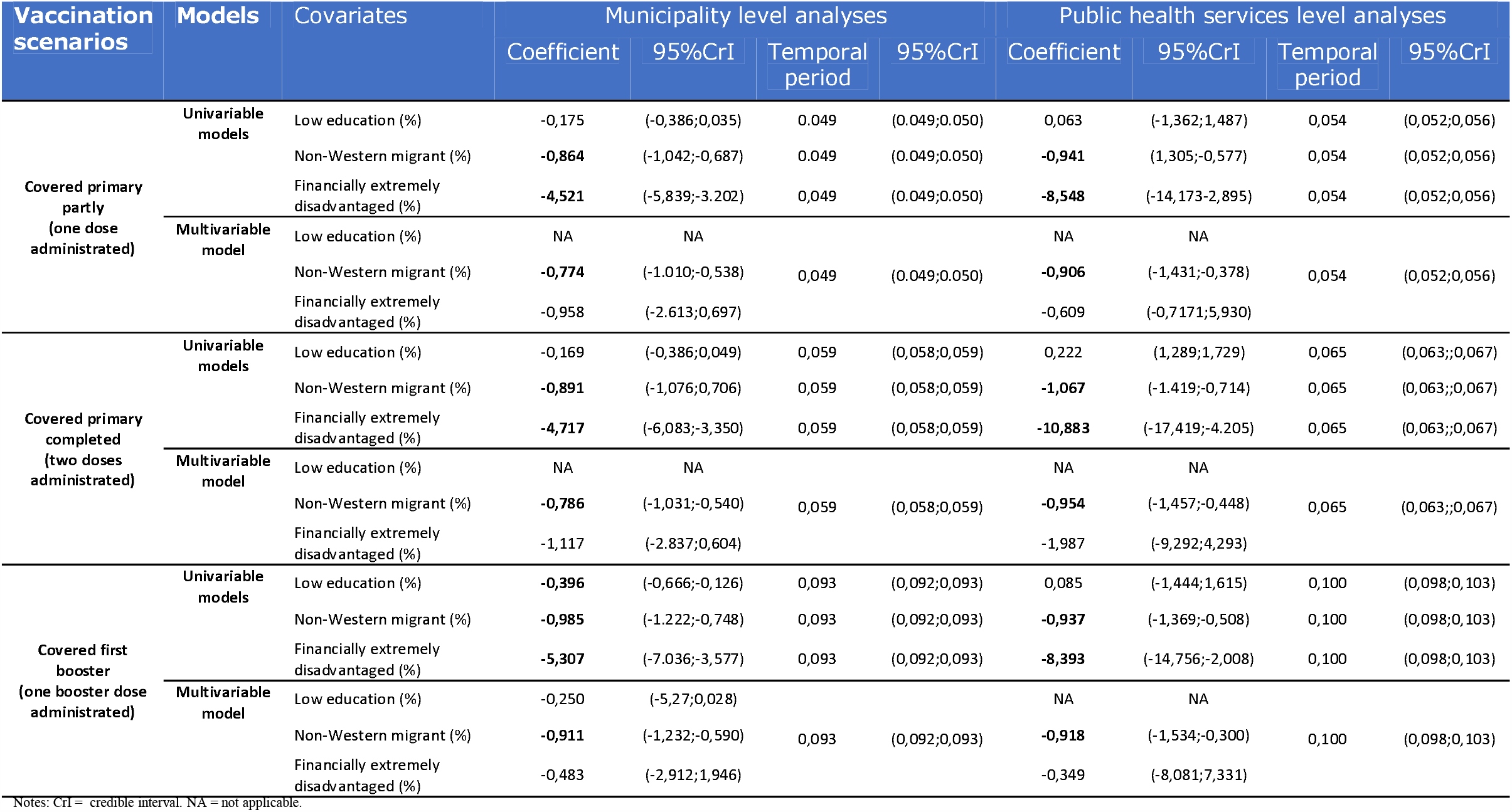
Model summary of Bayesian spatio-temporal ecological analysis of COVID-19 vaccination uptake in the Netherlands by vaccination scenarios.

## Discussion

This study explored the spatio-temporal distribution of the relative chance of COVID-19 vaccination uptake in the Netherlands with different COVID-19 vaccination scenarios ranging from covering primary partly (first dose) when the vaccines were just becoming available, over covering primary completed (second dose), to covering the first booster. We made use of the publicly available surveillance data of COVID-19 vaccination uptake which was routinely collected by RIVM. We applied Bayesian spatio-temporal modelling analysis for robust estimations accounting for spatial random effects and random noise across the spatio-temporal structure of the Netherlands on both the GGD and municipality levels. The known socio-demographic determinants of COVID-19 vaccination uptake were considered as socio-demographic spatial proxies to further fine-tune the estimates of the relative chance of vaccination uptake. As a result, COVID-19 campaigns and interventions can be better targeted to further close the vaccination gaps in the Netherlands.

### Spatio-temporal distribution of COVID-19 vaccination uptake

Overall, we observed a higher relative chance of COVID-19 vaccination uptake from the East and South of the Netherlands over the time periods included in this study for all three vaccination scenarios. The lowest relative chances of COVID-19 vaccination were identified in the main urban areas (in Dutch “Randstad”, which entails the agglomeration of cities in the west of the Netherlands, in particular, Amsterdam, Utrecht, Leiden, The Hague, and Rotterdam [35]) and the Bible belt region, i.e., the most religiously conservative region in the Netherlands.

It was within our expectation that people living in the Bible belt region would have a lower chance to take the COVID-19 vaccines, given the strong evidence of a lower chance of overall vaccination uptake in this region due to religions/believes [36-40]. Our findings thus confirmed a lower chance of COVID-19 vaccination uptake in this region, too. We suggest more efforts and behavioural interventions should be allocated to this region, such as attention to the engagement of trusted religious leaders and spokespeople [4, 41]. Also, a needs assessment to gather information regarding the problem should be conducted, which would probably lead to more effective interventions through an improved targeted public health communication [42].

However, surprisingly, we also found that people living in the Randstad region had a lower relative chance of vaccination uptake compared to the national average level. One reason for this finding may be the higher density of non-Western immigrants living in the Randstad area compared to the rest of the country, as reported by previous studies on the individual level in both the Netherlands [10, 11] and elsewhere [11, 15, 43] and summarised by a systematic review [44]. Our results from the ecological modelling analysis may also confirm this as a higher proportion of non-Western immigrants was associated with a lower relative chance of COVID-19 vaccination uptake on the populational level over time. Therefore, for future COVID-19 vaccination campaigns and interventions, more efforts may be needed toward this population. However, since “non-Western immigrants” is an umbrella term for different ethnicities and, possibly, different cultures, it might be necessary to specify the target population further when developing programs for vaccine hesitancy and tailor the programs according to the needs of the target population through needs assessments [10]. Another possible reason for this finding may be the potentially underestimated nature of our estimates. In the Netherlands, the national COVID-19 vaccination uptake surveillance only covers vaccines taken within the Netherlands but does not cover vaccines taken elsewhere. Given the relatively high proportion of Western immigrants living in the Randstad region as well [16], immigrants with a Western background may have taken the COVID-19 vaccines outside the Netherlands. This would lead to our estimations being underestimated overall.

When comparing the estimations between the GGD level and the municipality level, we demonstrated that refining the geographical scale can lead to enhanced insights. Different spatial patterns of the area with the lowest relative chance of COVID-19 vaccination uptake are sensitive to the choice of the spatial units, and different relative chances of COVID-19 vacation uptake among municipalities could be found within one GGD region. Take GGD-Zuid-Limburg as an example, a higher-than-average chance of COVID-19 vaccination uptake was observed in the Northwest municipalities over time, while municipalities located in the Southeast of GGD-Zuid-Limburg had a lower-than-average chance compared to the national level. This finding thus indicates the potential ecological fallacy in the ecological study. By aggregating data from a finer-defined geographic scale to a larger geographic scale, the internal heterogeneity within one larger geographic area, in our case municipalities nested within a GGD region, may not be captured and may lead to missing opportunities for public health actions and interventions. Given the fact that the GGDs are responsible for regional health, an increased need from certain municipalities, or even neighbourhoods, may not be noticed. We thus suggest that the GGDs should focus their monitoring strategies on a finer area level to identify potential public health problems early.

### Spatial socio-demographic proxies of COVID-19 vaccination uptake

In line with previous evidence which investigated how socio-demographic characteristics can influence one’s COVID-19 vaccination uptake on an individual level [13], our study confirmed that accounting for socio-demographic characteristics on the areal level can be helpful and could be applied as a spatial proxy for the COVID-19 vaccination uptake, too.

Throughout the COVID-19 scenarios (cover primary partially, cover primary completed and cover first booster), and on both GGD and municipality levels, we found that the proportion of non-Western immigrants was associated with a lower relative chance of COVID-19 vaccination uptake in the Netherlands. Our results are thus in line with the previous findings for the COVID-19 vaccination hesitancy [44].

Noteworthy is that the proportion of financially extremely disadvantaged individuals only played a significant negative association with COVID-19 vaccination cover primary completed scenario on both GGD and municipality levels, but not for other scenarios including covering primary partially and covering first booster in the multivariable models. Also, the proportion of people with a low level of education was not associated with vaccination uptake in any scenario on both geographic levels in the multivariable models. One of the reasons could also be that non-Western immigrants more often face financial difficulties and generally also often have a lower level of education in the Netherlands [45], therefore the impact of both might have disappeared due to adjusting for the proportion of non-Western immigrants on the population level. However, individual socio-demographic characteristics and their interactions with the environment are often assumed to play a bigger role in explaining health behaviours [20] and should therefore be considered in modelling above areal proxies whenever possible.

Also, the fact that our estimates for the impact of the spatial proxies differ/disappeared from the municipality level to the GGD level, indicated ecological fallacy, again, given that the spatial covariates across the municipalities provided more information compared to the GGD level.

Yet, our ecological findings still indicate that sub-populational needs from non-Western immigrants living in the Netherlands for additional COVID-19 vaccination services and efforts are not being met. Public health authorities and vaccination programmes could thus use our findings when designing future vaccination strategies for COVID-19 and other infectious diseases, such as mpox [46], by prioritising resources and services allocation and improving public health communication to regions and sub-populations that currently have a lower chance of vaccination uptake. This study thus helps in informing the (COVID-19-)vaccine-related intervention planners about whom and where to target in particular.

### Strength and limitations

The largest strength of this study is the application of Bayesian spatio-temporal modelling analysis on three different vaccination scenarios in the Netherlands over the crude prevalence calculation on two different geographical levels. As a result, this novel methodology produced more robust estimations of the posterior relative chance to identify low-risk clusters of COVID-19 vaccination uptake across different geographical units over the observed prevalence calculation on the small area level by accounting for the space-time autocorrelation using the weakly informative PC prior appointment [30, 47]. Another major strength of this study is the ecological assessment of socio-demographic characteristics relevant to COVID-19 vaccination on a small area level, to our knowledge, we were the first to explore such information in the Netherlands and provided a valuable recommendation for future vaccination strategies, which not only includes COVID-19 vaccination strategies but also for other future public health emergencies due to an infectious disease outbreak.

We acknowledge the following limitations of our study. First, we used the secondary surveillance data on COVID-19 vaccination uptake in the Netherlands, which was obligated to mask the proportion of vaccination when this was lower than five per cent. Our data for the spatial modelling analysis may thus be partly biased given the nature of our data. However, given the ability of the Bayesian spatio-temporal analysis we applied by accounting for the space-time autocorrelation [20, 48], we believe that such limitation would not majorly bias our smoothened estimations of the relative chance of the COVID-19 vaccination uptake [16, 20]. Second, our data only included the adult population living in the Netherlands and excluded the population younger than 18-years-old, due to the COVID-19 vaccines being only approved for minors at a much later stage in Europe and the Netherlands [29, 49]. Given the fact that these minors are equally important in COVID-19 prevention, and their COVID-19 vaccination uptake can be largely influenced by their guardian’s beliefs on vaccination, our findings may not be able to be generalised in this population, and hence future studies may need to focus on this population, such as guardians’ vaccination status and attitudes. Third, previous studies have shown that specific immigrant groups, such as Turkish immigrants, showed higher COVID-19 vaccination hesitancy in Europe [15], as well as a higher cumulative incidence of COVID-19 infections, such as Ghanaian, Moroccan, Turkish, and African Surinamese living in the Netherlands [11]. Due to the lack of data on the proportion of specific ethnic groups, our data did not distinguish between specific ethnic groups of migrants and, thus, we used an aggregated variable “non-Western migrant”, which may have failed to account for ethnic clusters. Future studies should therefore investigate the ethnic clusters when accurate data are available, to better tailor vaccination and other interventions deliveries. Fourth, our data did not include information on the uptake of specific types of COVID-19 vaccines and hence could not identify the different vaccines’ clusters spatially and temporally with a subgroup analysis. Given the established effectiveness of different vaccines against different COVID-19 variants [50], such information can be useful and should be investigated in the future to serve as improved evidence for advanced mathematical modelling on the COVID-19 pandemic.

### Conclusions

In conclusion, we showed that the relative chance of COVID-19 vaccination uptake was heterogenous for all three rounds of vaccination scenarios over time in the Netherlands on both GGD and municipality levels. Estimations on the municipality level show vaccination variability and more concise clustering patterns compared to the GGD level, and thus provide more insights into the strategies for vaccination services. We identified regions, such as the Bible belt region, and regions with a higher proportion of non-Western immigrants, that had a lower chance of COVID-19 vaccination uptake. In the transitioning era towards the COVID-19 endemic, our results should inform public health professionals that more efforts are needed to reach individuals living in these regions in the Netherlands for the ongoing second COVID-19 booster campaign to prevent more serious COVID-19-related health outcomes. Our findings can also provide valuable information for other infectious diseases to further close the vaccination gaps in general.

## Data Availability

All data produced in the present study are available upon reasonable request to the authors, and available at https://coronadashboard.government.nl/landelijk/vaccinaties

## Notes

### Competing Interest Statement

The authors have declared no competing interest.

### Funding Statement

This study did not receive any funding

