## Supplementary file for "Spatio-temporal distributions of COVID-19 vaccine doses uptake in the Netherlands: A Bayesian ecological modelling analysis"

### Supplementary materials

Haoyi Wang^1,2§^, Tugce Varol^1^, Thomas Gültzow^1^, Hanne M. L. Zimmermann^1,3^, Robert A.C. Ruiter^1^, Kai J. Jonas^1^

^1^Department of Work and Social Psychology, Maastricht University, Maastricht, The Netherlands

^2^Viroscience Department, Erasmus Medical Centre, Rotterdam, The Netherlands

^3^Department of Infectious Diseases, Public Health Service of Amsterdam, Amsterdam, The Netherlands

^§^Correspondence to: Haoyi Wang

Department of Work and Social Psychology, Maastricht University, Maastricht, 6200ER, the Netherlands

#### S1 Spatio-temporal distribution of the COVID-19 uptake covered primary partly on the municipality level

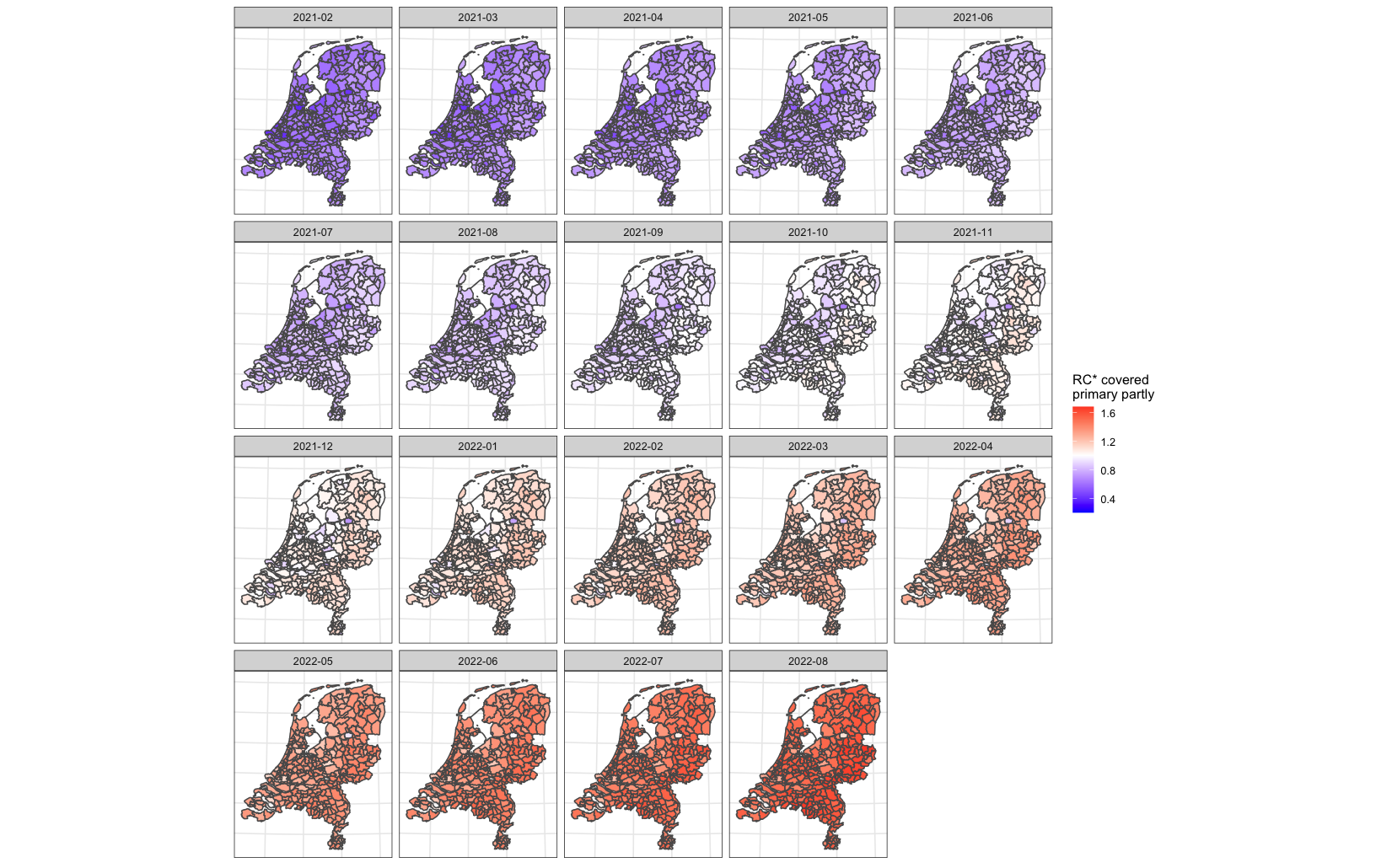

#### S2 Spatio-temporal distribution of the COVID-19 uptake covered primary partly on the public health services (GGD) level

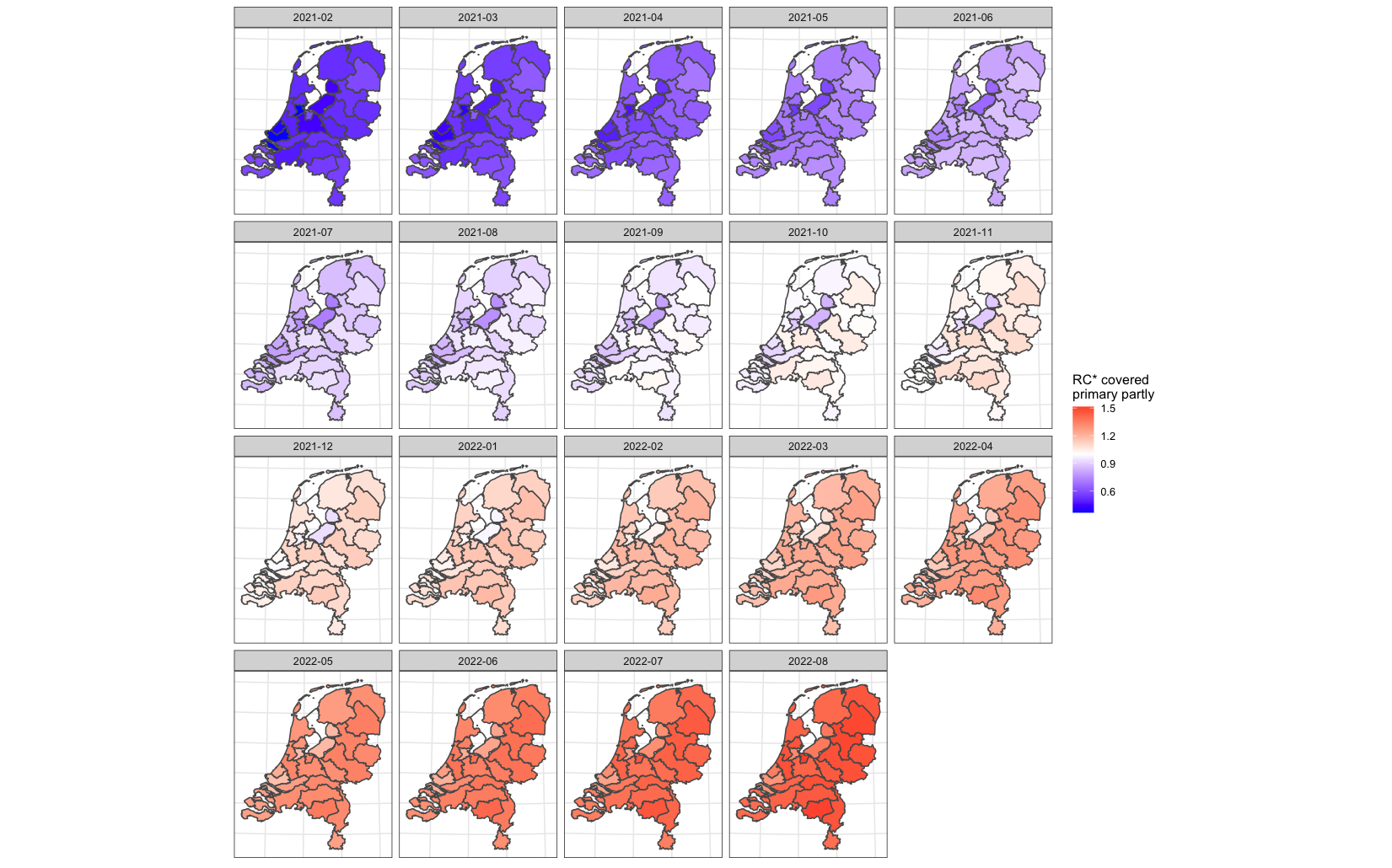

#### S3 Spatio-temporal distribution of the COVID-19 uptake covered primary completed on the municipality level

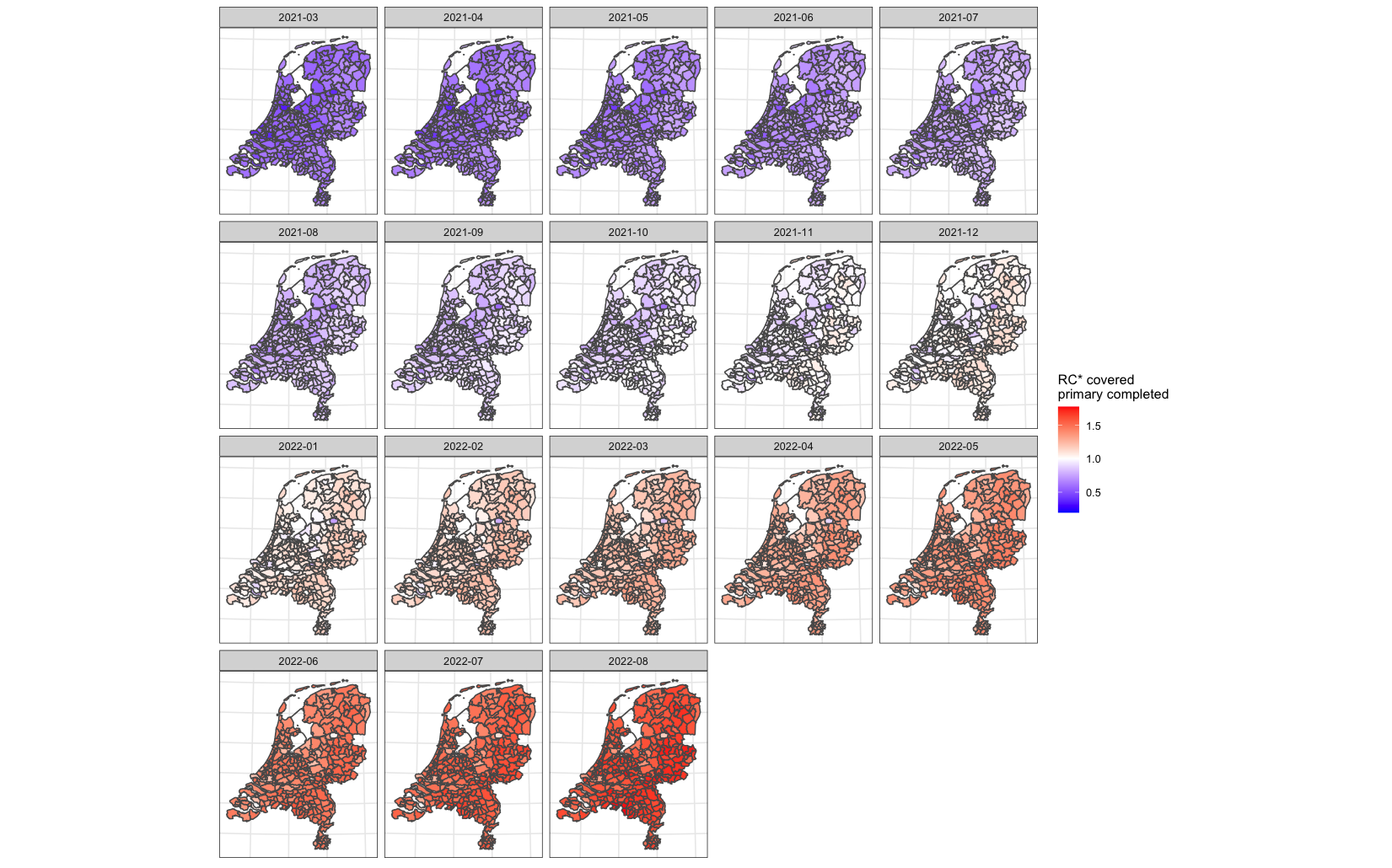

#### S4 Spatio-temporal distribution of the COVID-19 uptake covered primary completed on the public health services (GGD) level

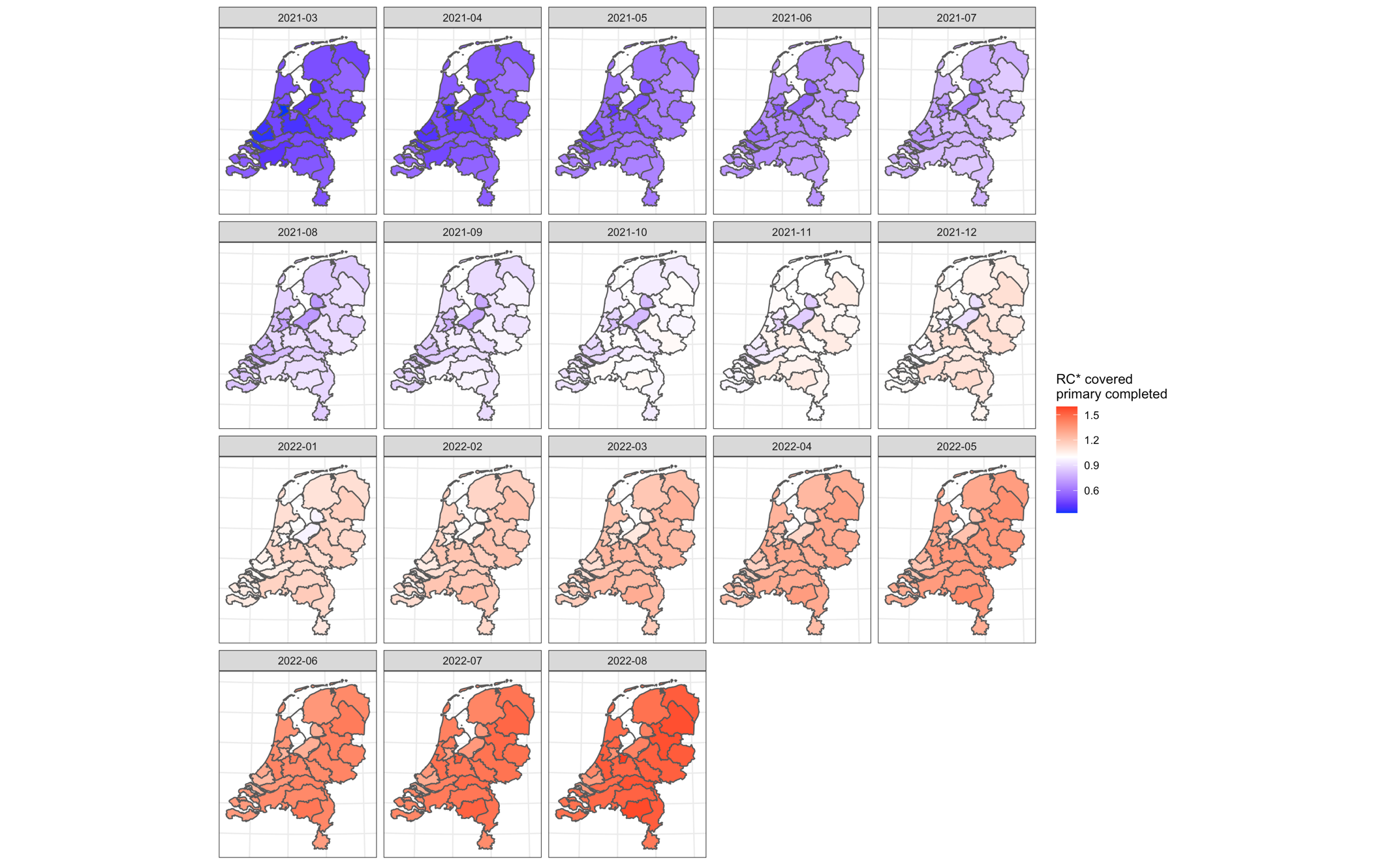

#### S5 Spatio-temporal distribution of the COVID-19 uptake covered first booster on the municipality level

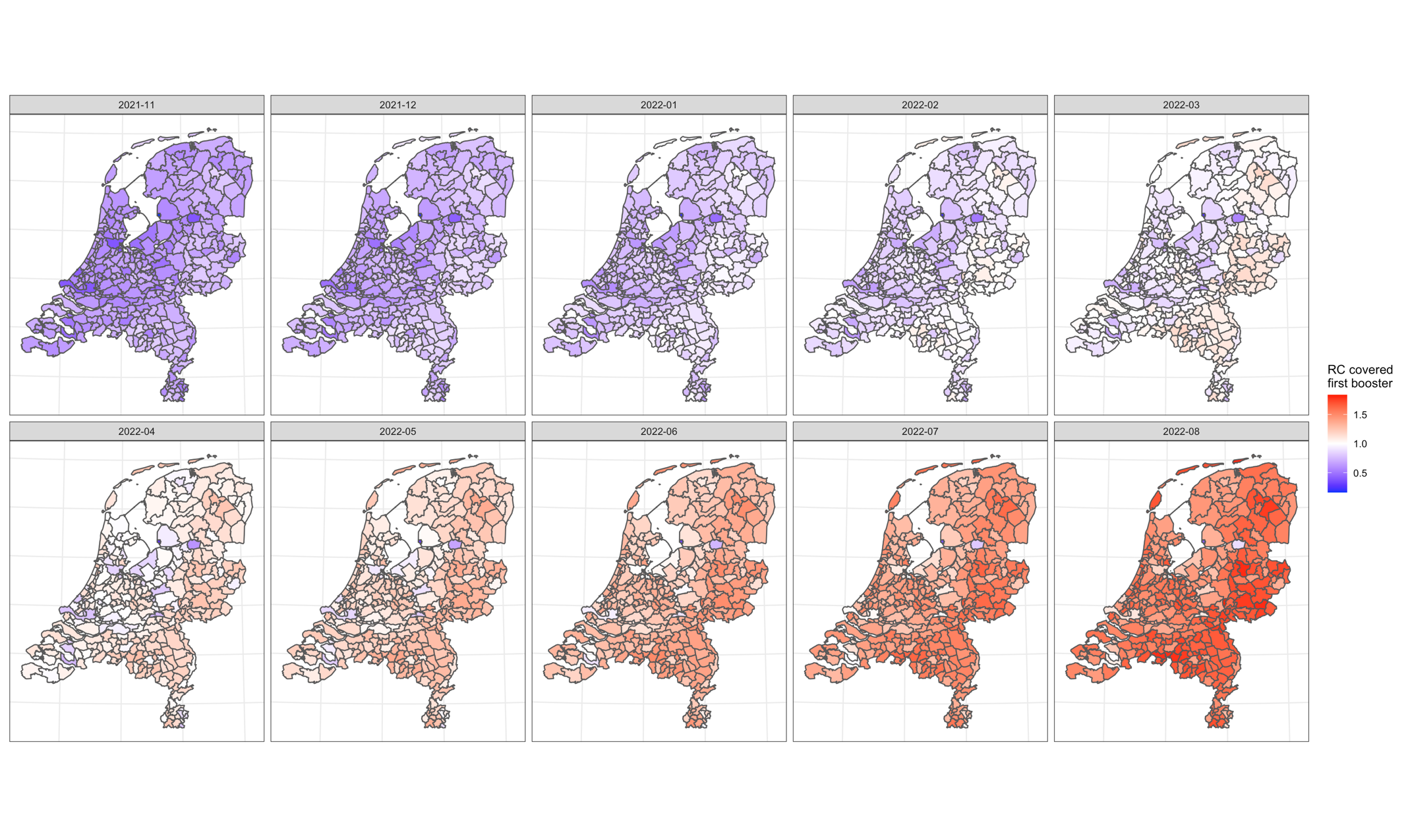

#### S6 Spatio-temporal distribution of the COVID-19 uptake covered first booster on the public health services (GGD) level

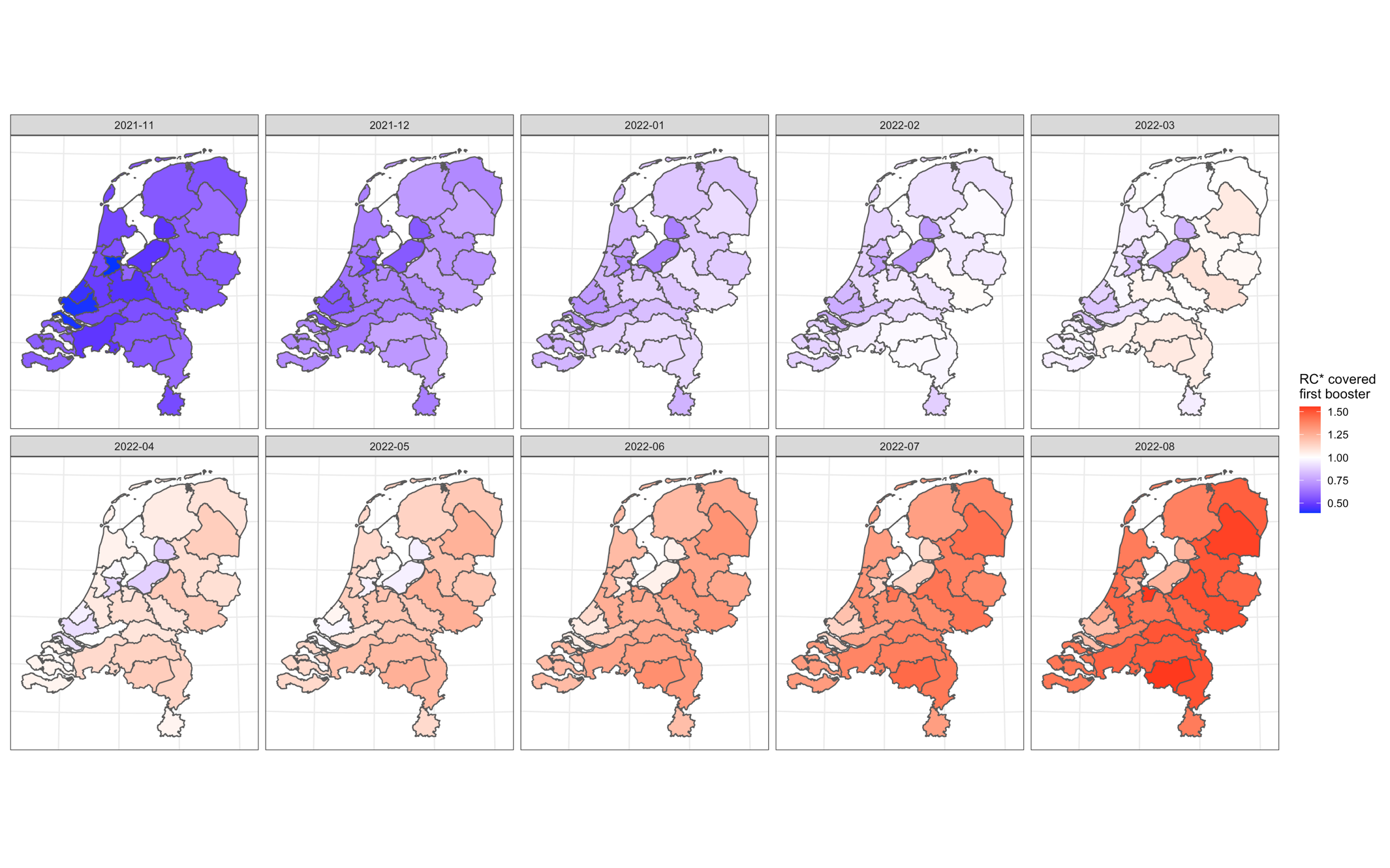

#### S7 Spatial distribution of the COVID-19 uptake covered primary partly on the municipality level over the selected periods

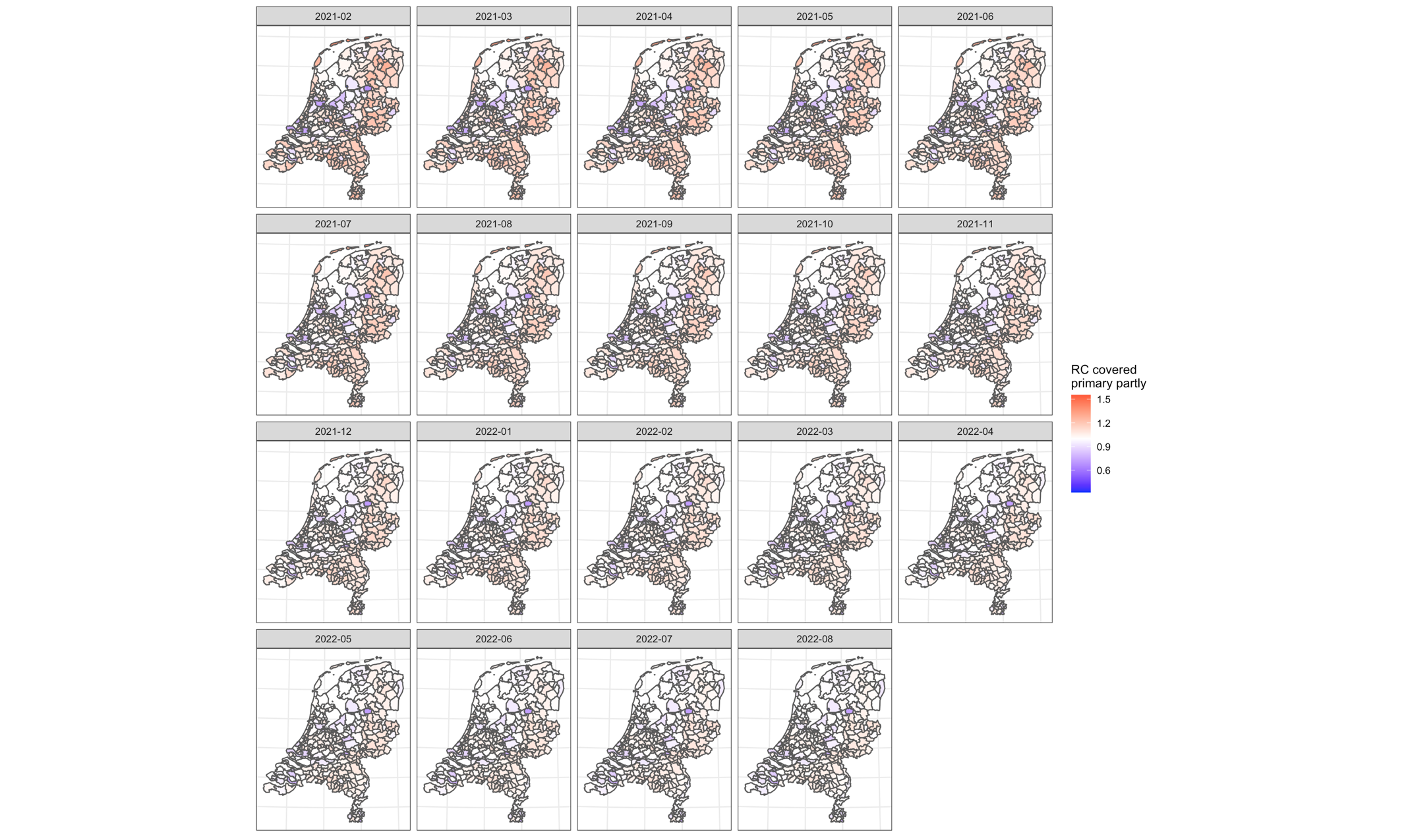

#### S8 Spatial distribution of the COVID-19 uptake covered primary partly on the public health services (GGD) level over the selected periods

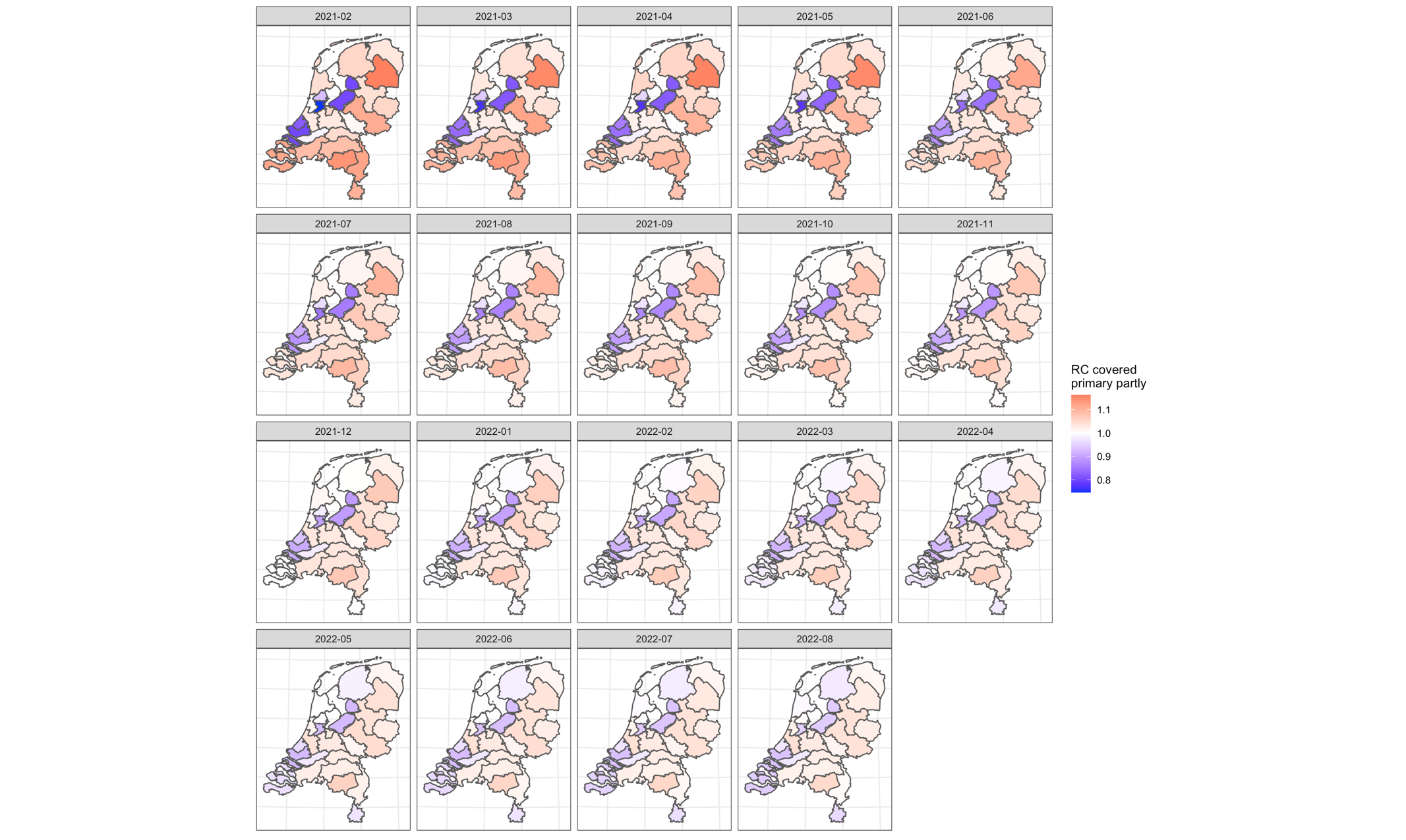

#### S9 Spatial distribution of the COVID-19 uptake covered primary completed on the municipality level over the selected periods

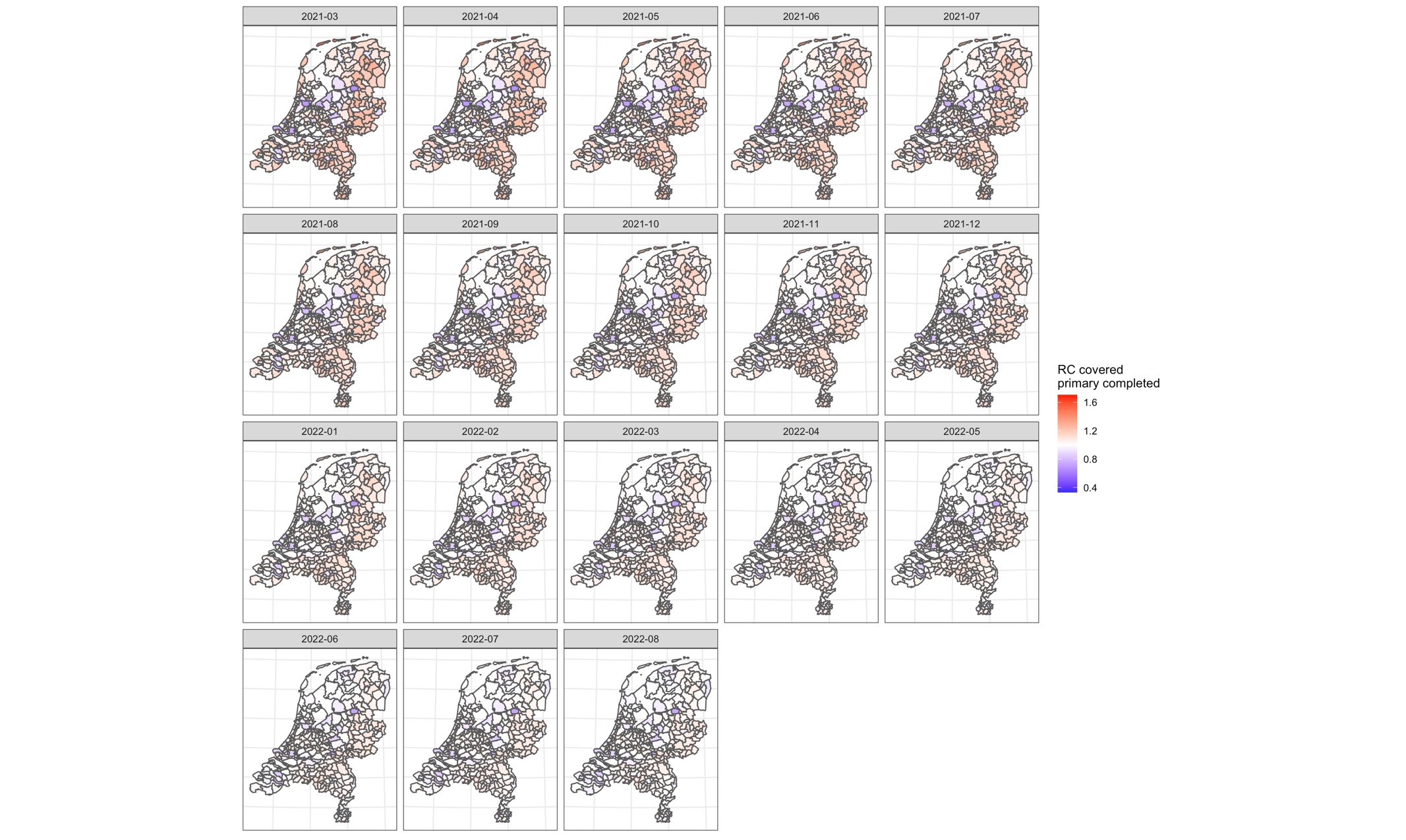

#### S10 Spatial distribution of the COVID-19 uptake covered primary completed on the public health services (GGD) level over the selected periods

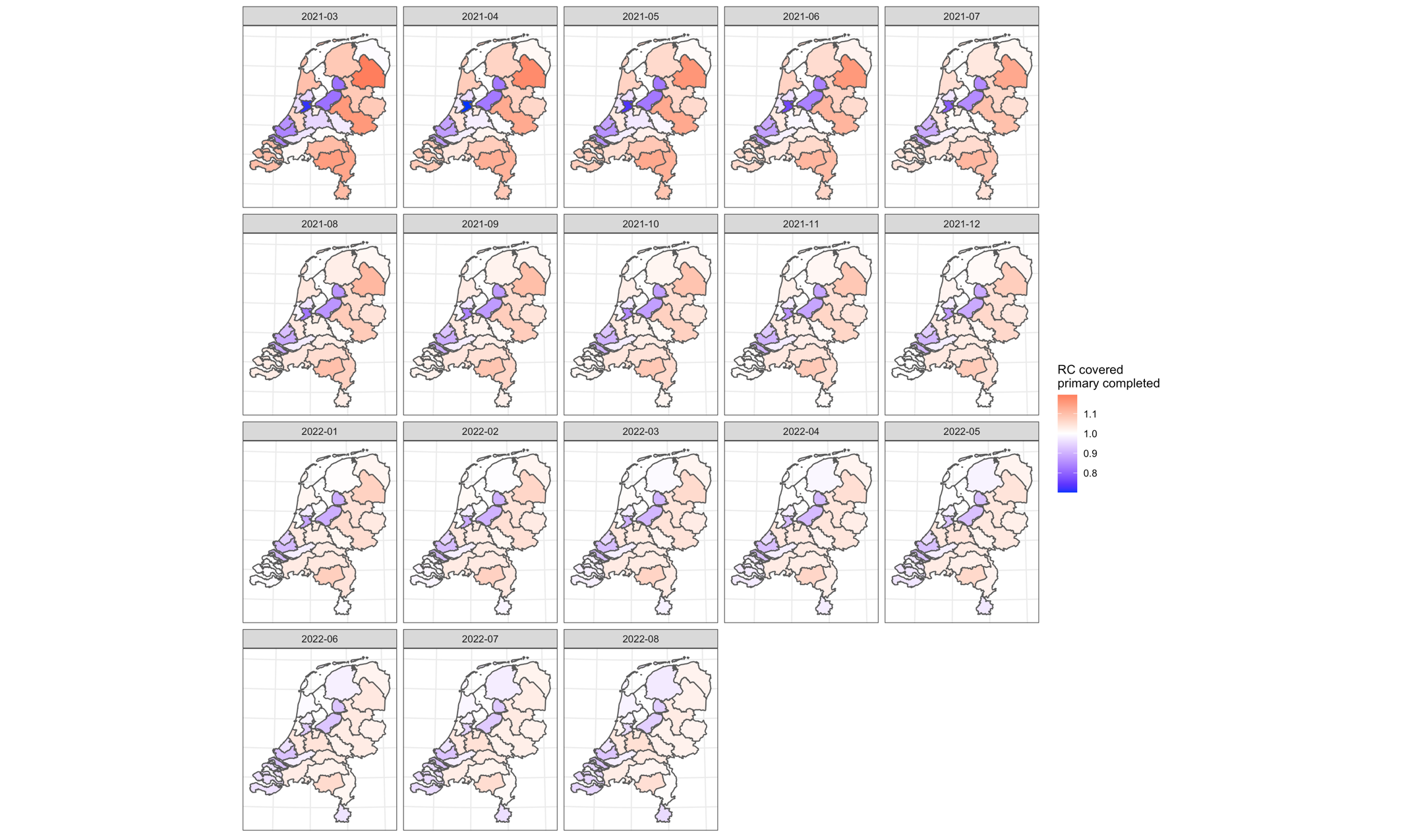

#### S11 Spatial distribution of the COVID-19 uptake covered first booster on the municipality level over the selected periods

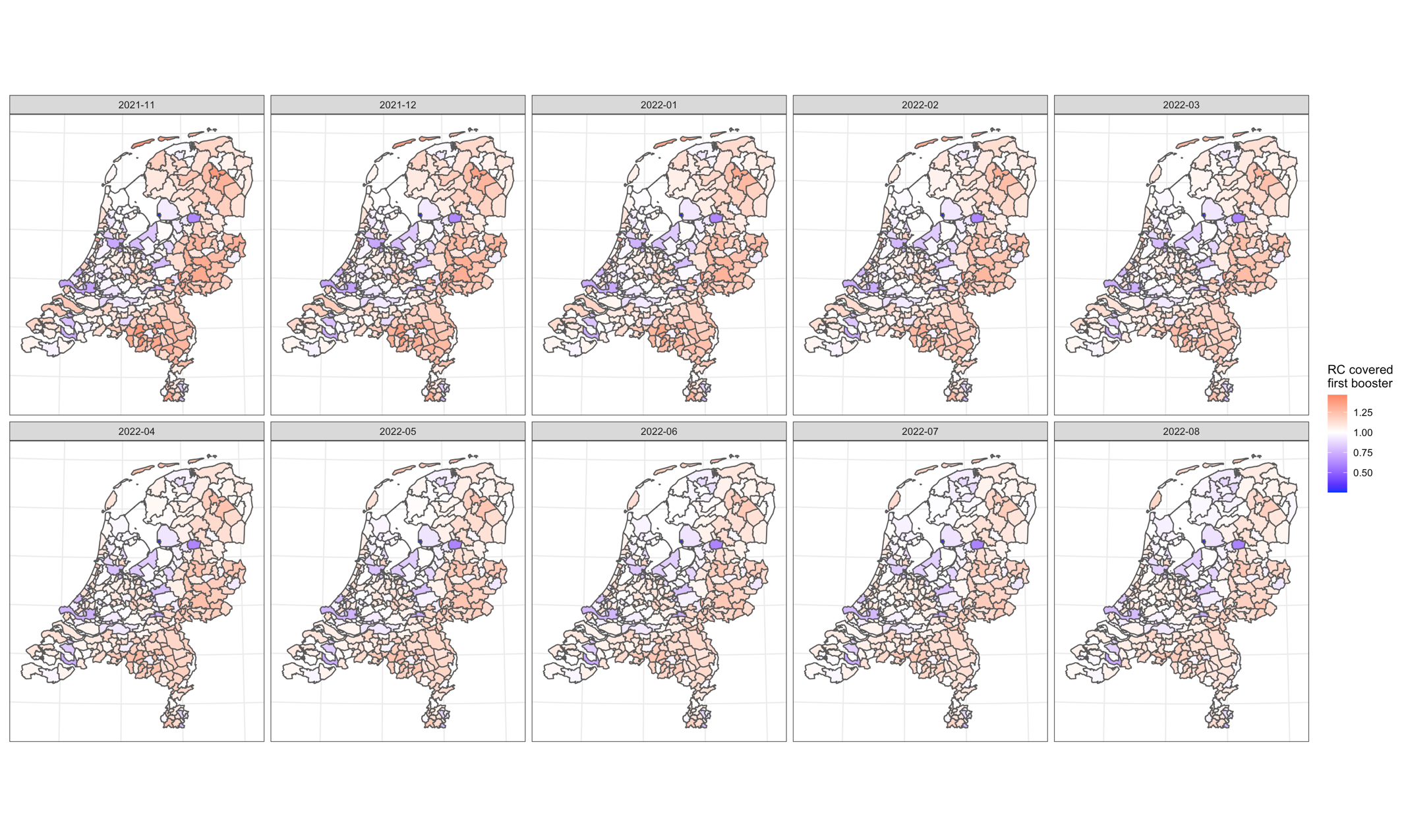

#### S10 Spatial distribution of the COVID-19 uptake covered first booster on the public health services (GGD) level over the selected periods

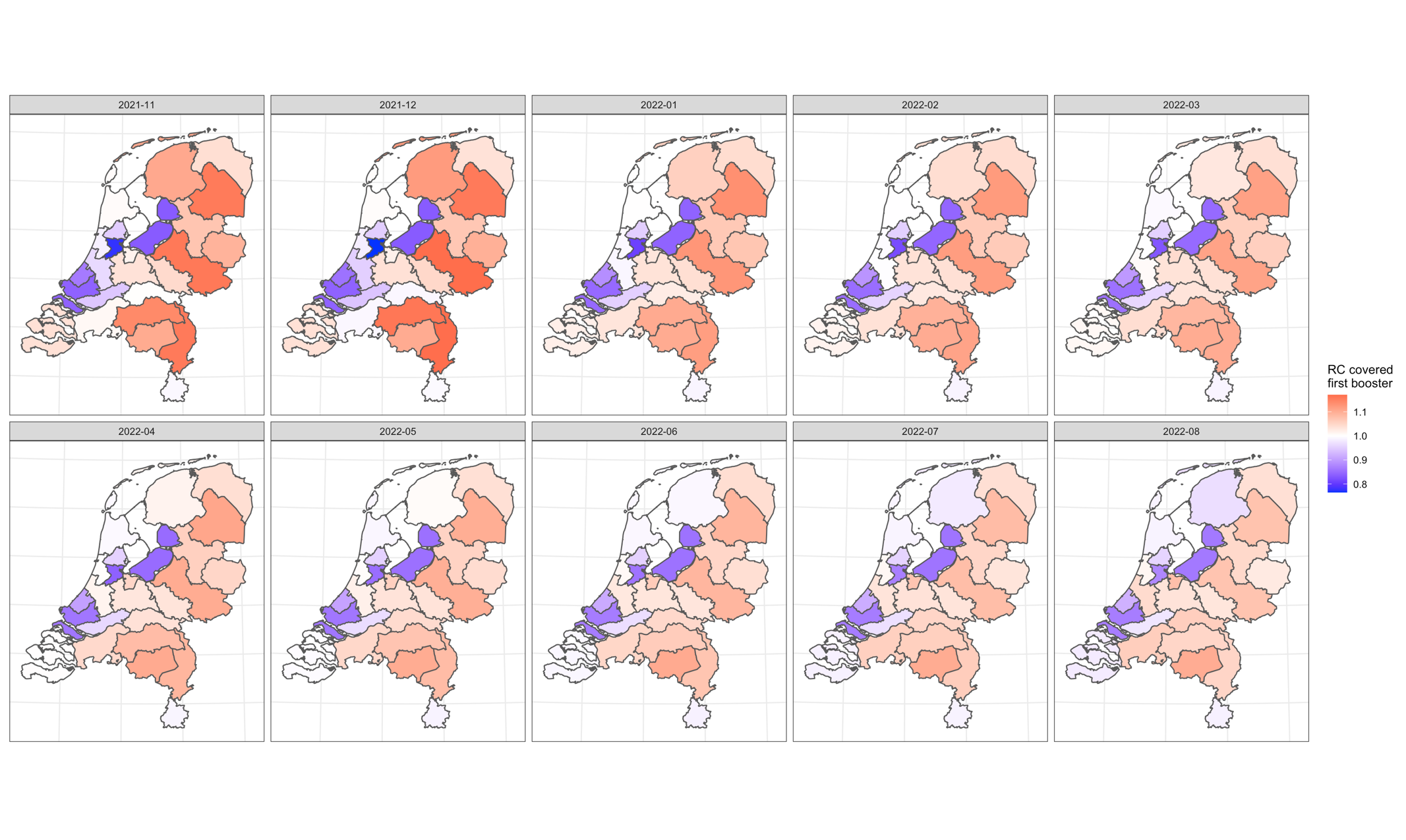
